# *ERBB4* Mutant Alleles Found in *BRAF* WT Melanomas That Drive the Proliferation of a *BRAF* WT Melanoma Cell Line

**DOI:** 10.1101/2022.06.21.22276707

**Authors:** Lauren M. Lucas, Richard L. Cullum, Joelle N. Woggerman, Vipasha Dwivedi, Jessica A. Markham, Connor M. Kelley, Elizabeth L. Knerr, Laura J. Cook, Megan A. Jacobi, Darby C. Taylor, Cristina C. Rael, Howard C. Lucas, Damien S. Waits, Taraswi M. Ghosh, Kenneth M. Halanych, Ram B. Gupta, David J. Riese

## Abstract

Metastatic skin cutaneous melanomas that contain wild-type *BRAF* alleles (“*BRAF* WT melanomas”) remain a significant clinical challenge, primarily because of the paucity of targets for therapeutic intervention. In prior work, *in silico* analyses of The Cancer Genome Atlas Skin Cutaneous Melanoma (TCGA-SKCM) dataset suggested that elevated transcription of the gene that encodes the ERBB4 receptor tyrosine kinase may drive *BRAF* WT melanomas. Moreover, that prior work demonstrated that expression of the wild-type ERBB4 gene (WT *ERBB4*) stimulates clonogenic proliferation by the IPC-298, MEL-JUSO, MeWo, and SK-MEL-2 human *BRAF* WT melanoma cell lines. Moreover, expression of a dominant-negative (K751M) *ERBB4* mutant (*ERBB4* DN) inhibits clonogenic proliferation by these same cell lines.

Here we have extended these findings by investigating the role of *ERBB4* mutant alleles in *BRAF* WT melanomas. *In silico* analyses of the TCGA-SKCM *BRAF* WT melanoma dataset indicates that *ERBB4* missense mutant alleles occur in a non-random manner, suggesting that melanomagenesis selects for the *ERBB4* missense mutant alleles. Specifically, *ERBB4* missense mutant alleles affect amino acid residues that are weakly correlated with residues conserved in the ERBB3 extracellular domains and the EGFR tyrosine kinase domain. The occurrence of *ERBB4* missense mutant alleles in the TCGA-SKCM *BRAF* WT melanoma dataset is weakly inversely correlated with events that cause ERBB4-independent PI3K pathway signaling and is strongly correlated with events that cause elevated RAS pathway signaling. Thus, the *in silico* analyses suggest that *ERBB4* mutant alleles encode proteins that stimulate PI3K signaling, which cooperates with elevated RAS signaling to drive *BRAF* WT melanomas. Moreover, the *in silico* analyses have prioritized the *ERBB4* mutant alleles as candidate drivers of *BRAF* WT melanomas. Two *ERBB4* mutant alleles (G85S and G741E) found in *BRAF* WT melanomas stimulate clonogenic proliferation of MEL-JUSO *BRAF* WT melanoma cells. We discuss these findings in the context of strategies for identifying and treating *ERBB4*-dependent *BRAF* WT melanomas.

## Introduction

BRAF inhibitors, MEK inhibitors, and immune checkpoint inhibitors (“checkpoint inhibitors”) are transforming the treatment of advanced skin cutaneous melanomas that possess oncogenic *BRAF* mutations (“*BRAF* mutant melanomas”). The Surveillance, Epidemiology, and End Results Program of the National Cancer Institute [1] reports a 25% five-year survival of metastatic skin cutaneous melanoma patients for the period of 2010 to 2018 [2]. In contrast, a 2019 clinical trial reports 34% five-year survival of patients with advanced *BRAF* mutant skin cutaneous melanomas treated with BRAF and MEK inhibitors [3, 4]. A 2019 clinical trial reports 60% five-year survival of patients with advanced *BRAF* V600X mutant skin cutaneous melanomas *(“BRAF* V600X mutant melanomas”) treated with a combination of immune checkpoint inhibitors [3, 5]. Finally, combining immune checkpoint inhibitors with BRAF and MEK inhibitors will likely lead to further improvements in survival [6].

Unfortunately, approximately 50% of advanced skin cutaneous melanomas possess wild-type *BRAF* alleles, and contemporary treatments of advanced skin cutaneous melanomas that contain wild-type *BRAF* (“*BRAF* WT melanomas”) have yielded less impressive results. In part, these less impressive results are because of a paucity of actionable targets for the (targeted) treatment of these tumors [7]. Moreover, the five-year survival of patients with advanced *BRAF* WT skin cutaneous melanomas treated with immune checkpoint inhibitors is only 48%, less than the 60% experienced by patients with advanced *BRAF* mutant skin cutaneous melanomas in a parallel study [5]. Hence, our previous work attempted to address this gap in treatment efficacy by evaluating the ERBB4 receptor tyrosine kinase as a candidate target in *BRAF* WT skin cutaneous melanomas [8].

ERBB4 (HER4) is a member of the ERBB family of receptor tyrosine kinases (RTKs), which includes the epidermal growth factor receptor (EGFR), ERBB2 (HER2/Neu), and ERBB3 (HER3). ERBB4 possesses extracellular ligand-binding domains, a single-pass hydrophobic transmembrane domain, an intracellular tyrosine kinase domain, and intracellular tyrosine residues that function as phosphorylation sites (**Figure 1**). Ligand binding to EGFR, ERBB3, or ERBB4 stabilizes the receptor extracellular domains in an open conformation competent for symmetrical homodimerization and heterodimerization of the receptor extracellular domains. The dimerization of the extracellular domains enables asymmetrical dimerization of the receptor cytoplasmic domains. Phosphorylation of one receptor monomer on tyrosine residues by the tyrosine kinase domain of the other receptor monomer (“cross-phosphorylation”) ensues (**Figure 2**). This tyrosine phosphorylation creates binding sites for effector proteins and activation of downstream signaling pathways [9].

**Figure 1.**
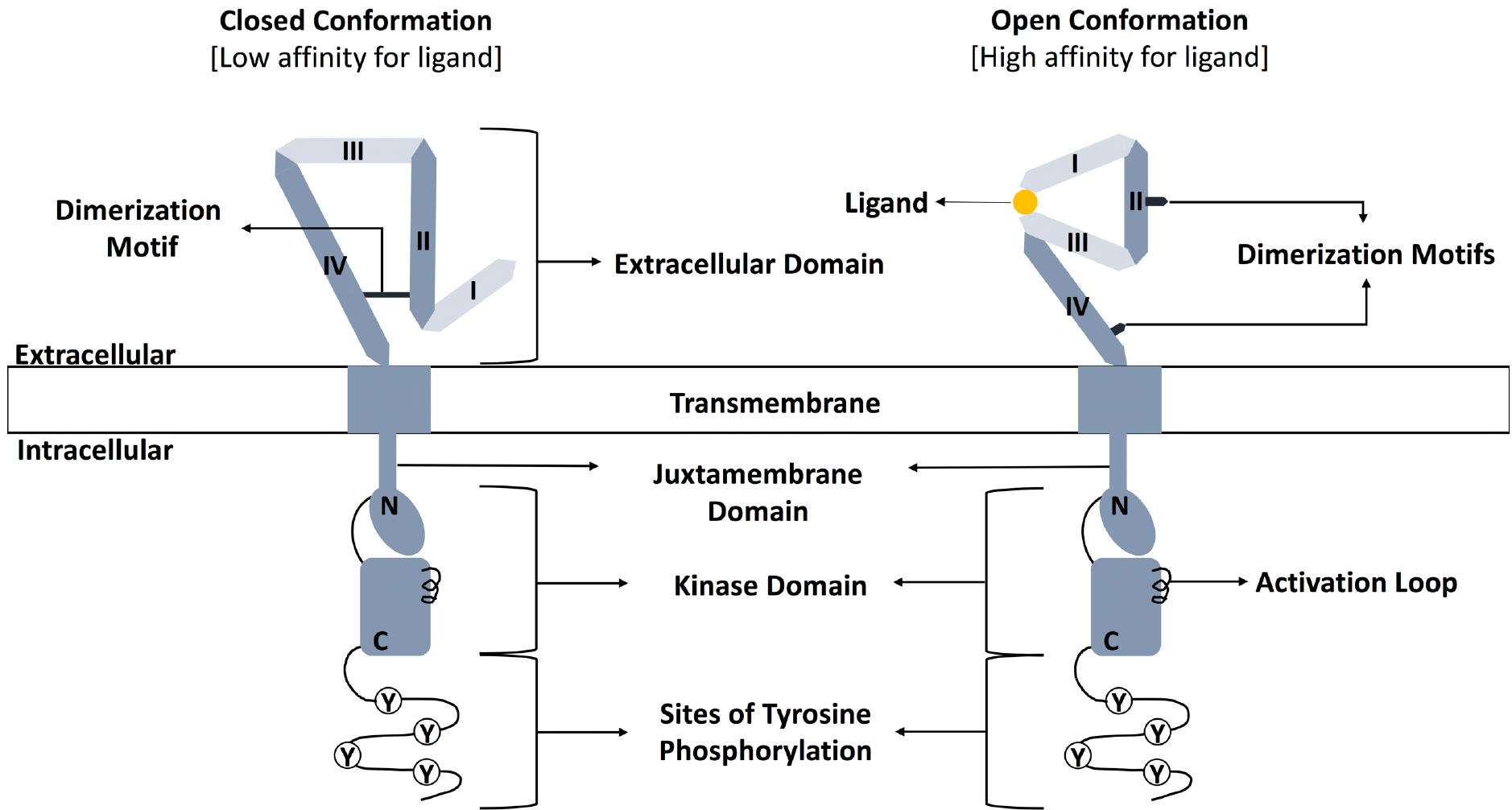
The extracellular domains of ERBB4 exist in an equilibrium between the closed conformation that has low affinity for ligand and buried dimerization motifs and the open conformation that has high affinity for ligand and has exposed dimerization motifs. Adapted from [9].

**Figure 2.**
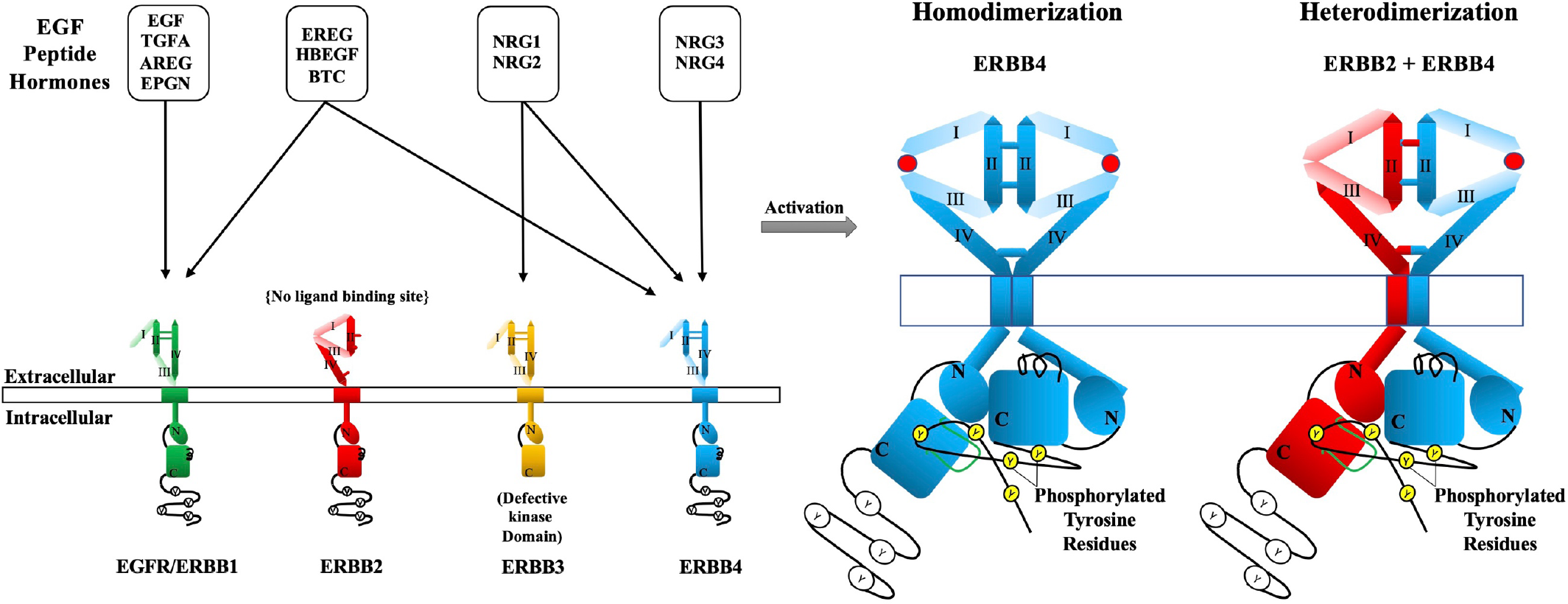
ERBB4 ligands stimulate ERBB receptor signaling via ERBB4 homodimerization and heterodimerization. Adapted from [9].

Elevated signaling by an RTK is a hallmark of many types of cancer. Hence, RTK overexpression, ligand overexpression, and gain-of-function mutations in an RTK gene are all mechanisms for pathologic, elevated RTK signaling [10]. Indeed, EGFR and ERBB2 have been validated as targets for therapeutic intervention in numerous types of tumors; monoclonal antibodies and small molecular tyrosine kinase inhibitors have been approved to treat tumors dependent on these receptors [11–26]. It appears that ERBB3, particularly ERBB3-ERBB2 heterodimers, also drives various human tumors [27, 28].

In contrast, the role that ERBB4 plays in human tumors remains ambiguous. Part of the ambiguity reflects that an ERBB4 homodimer can function as a tumor suppressor, whereas an ERBB4-EGFR or ERBB4-ERBB2 heterodimer can drive tumor cell proliferation or aggressiveness [9]. Our previous work has demonstrated that ectopic expression of wild-type *ERBB4* stimulates the proliferation of *BRAF* WT melanoma cell lines and ectopic expression of a dominant-negative (K751M) mutant *ERBB4* allele inhibits the proliferation of *BRAF* WT melanoma cell lines. Genes that encode ligands for ERBB4 are transcribed by these cell lines. Moreover, ectopic expression of wild-type *ERBB2* stimulates the proliferation of a *BRAF* WT melanoma cell line and ectopic expression of a dominant-negative (K753A) mutant *ERBB2* allele inhibits the proliferation of a *BRAF* WT melanoma cell line. These data suggest that ligand-induced signaling by ERBB4-ERBB4 heterodimers drive *BRAF* WT melanomas [8].

Structural studies of ERBB family receptors and analyses of *ERBB* mutant alleles have revealed multiple mechanisms by which a missense mutation in an *ERBB* gene may cause elevated (oncogenic) signaling by that receptor protein [29]. (1) A mutation may increase the affinity of a receptor for its cognate ligands. (2) A mutation may cause a shift in the extracellular domain conformational equilibrium; the mutant may now favor the open conformation (which is competent for dimerization and signaling) over the closed conformation (which is not competent for dimerization and signaling). (3) A mutation may directly increase receptor dimerization, through the creation of novel intermolecular chemical bonds. Such activating mutations may affect dimerization of the extracellular domains, transmembrane domain, or intracellular kinase domain. (4) A mutation may destabilize the inactive conformation of the kinase domain or stabilize the active conformation of the kinase domain. (5) The substitution of a glutamate residue for a tyrosine residue at the site of tyrosine phosphorylation may mimic phosphorylation, resulting in constitutive binding of effector proteins and activation of downstream signaling events.

*ERBB4* mutant alleles, most of which remain largely uncharacterized, have been found in several cancers [9]. A previous report suggested that *ERBB4* mutant alleles found in melanoma samples drive the proliferation of melanomas [30]. However, this finding has neither been replicated nor extended [9]. One potential explanation is that this work and subsequent efforts failed to account for the possibility that ERBB4 homodimers are growth inhibitory. Another possibility is that this work employed proprietary cell lines and focused on both *BRAF* V600X and WT melanomas [30].

To address these issues, we have previously identified four commercially-available, ERBB4-dependent *BRAF* WT human melanoma cell lines (IPC-298, MEL-JUSO, MeWo, and SK-MEL-2) [8]. Here we identify *ERBB4* mutant alleles present in *BRAF* WT human melanoma samples and we have used an *in silico* workflow to prioritize those mutant alleles that are the most likely to function as drivers of *BRAF* WT melanomas. Finally, we have used the commercially-available, ERBB4-dependent MEL-JUSO *BRAF* WT melanoma cell line to identify the *ERBB4* mutant alleles that stimulate the clonogenic proliferation of these cells.

## Results

### A. ERBB4 non-synonymous mutations occur at a non-stochastic frequency

Our *in silico* analysis of *ERBB4* mutant alleles is based on The Cancer Genome Atlas Skin Cutaneous Melanoma (TCGA-SKCM) dataset (470 cases) [31]. Most of the analyses reported here focus on the 227 cases that possess wild-type (WT) *BRAF* alleles (“*BRAF* WT melanomas”) [8, 31].

In the TCGA-SKCM dataset, *ERBB4* missense mutant alleles are slightly more common in *BRAF* WT melanomas than *BRAF* V600X mutant melanomas (15.4% and 11.4% respectively). Therefore, we hypothesized that *ERBB4* mutant alleles are selected in *BRAF* WT melanomas, thereby justifying the analyses reported in this work.

The ratio of non-synonymous (missense, stop-gained, and frameshift) to synonymous (synonymous, stop-retained) mutations is an indicator of selection for mutant alleles by tumor cells. Generally speaking, if the ratio of non-synonymous to synonymous mutant alleles (N/S ratio) in a particular gene is greater than 1, it is indicative of selection for non-synonymous mutant alleles in that gene [32].

Because of the exposure of skin cells to ultraviolet (UV) light, melanomas carry numerous cytosine to thymine mutations, many of which function as passengers. This precludes using an N/S cutoff of 1 to identify genes for which mutant alleles function as drivers [33]. Thus, we have compared the N/S ratio for *ERBB4* in the 227 *BRAF* WT melanomas of the TCGA-SKCM dataset against the N/S ratio for genes (*ARMC4, CFTR, ERC2, SLIT2,* and *SLIT3)* that are roughly the same size as *ERBB4* and that from the literature do not appear to function as melanoma drivers.

The N/S ratio for *ERBB4* is 5.33, which is significantly greater (p=0.003) than the average N/S ratio for the five control genes (2.44) (**Table 1**). Moreover, the N/S ratio for the *KRAS* and *NRAS* oncogenes and the *CDKN2A*, *NF1*, *RB*, and *TP53* tumor suppressor genes is also greater than the average N/S ratio for the five control genes. In contrast, the N/S ratio for EGFR, ERBB2, and ERBB3 genes is not significantly greater than the average N/S ratio for the five control genes. These results suggest that *ERBB4* mutant alleles, but not *EGFR*, *ERBB2*, or *ERBB3* mutant alleles, function as drivers in *BRAF* WT melanomas.

**Table 1.** The ratio of non-synonymous to synonymous (N/S ratio) *ERBB4* mutations in the TCGA-SKCM *BRAF* WT melanoma dataset is significantly greater than the average N/S ratio in the *ARMC4, CFTR, ERC2, SLIT2,* and *SLIT3* genes.

|  | Non-synonymous | Synonymous | N/S Ratio |  |
| --- | --- | --- | --- | --- |
|  |  |  | Mean | Std Dev |
| **Control Genes** |  |  | 2.44 | 1.23 |
| *ARMC4* | 60 | 13 | 4.62 |  |
| *CFTR* | 37 | 18 | 2.06 |  |
| *ERC2* | 53 | 28 | 1.89 |  |
| *SLIT2* | 50 | 26 | 1.92 |  |
| *SLIT3* | 49 | 29 | 1.69 |  |
| **ERBB4 Gene** |  |  | N/S Ratio | One Sample T-test Against Control Genes |
| *ERBB4* | 48 | 9 | 5.33 | p = 0.003 |
| **Oncogenes** |  |  | N/S Ratio |  |
| *KRAS* | 12 | 1 | 12.00 |  |
| *NRAS* | 126 | 1 | 126.00 |  |
| **Tumor Suppressor Genes** |  |  | N/S Ratio |  |
| *CDKN2A* | 25 | 6 | 4.17 |  |
| *NF1* | 59 | 15 | 3.93 |  |
| *RB1* | 13 | 2 | 6.50 |  |
| *TP53* | 35 | 3 | 11.67 |  |
| **ERBB1-3 Genes** |  |  | N/S Ratio |  |
| *EGFR* | 25 | 10 | 2.50 |  |
| *ERBB2* | 5 | 2 | 2.50 |  |
| *ERBB3* | 6 | 5 | 1.20 |  |

### B. ERBB4 mutations are somewhat more likely to affect functionally conserved amino acid residues

Using the TCGA-SKCM, *BRAF* WT melanoma dataset, we identified 40 *ERBB4* missense mutations that are not coincident with an *ERBB4* stop-gained mutation or splice site mutation; one of these *ERBB4* mutant alleles (R711C) was found in three tumor samples and the other 39 *ERBB4* mutant alleles were found in a single tumor sample. Moreover, *ERBB4* mutant alleles affect every functional region of ERBB4: extracellular subdomain I, extracellular subdomain II, extracellular subdomain III, extracellular subdomain IV, the transmembrane domain (T), and the tyrosine kinase domain (K) (**Figure 3**). The single mutational hotspot (R711C) is assigned a priority point to help identify the best candidates for *ERBB4* driver mutant alleles in *BRAF* WT melanomas. However, we employed other approaches to prioritize candidate *ERBB4* mutant alleles that are likely to function as *BRAF* WT drivers.

**Figure 3.**
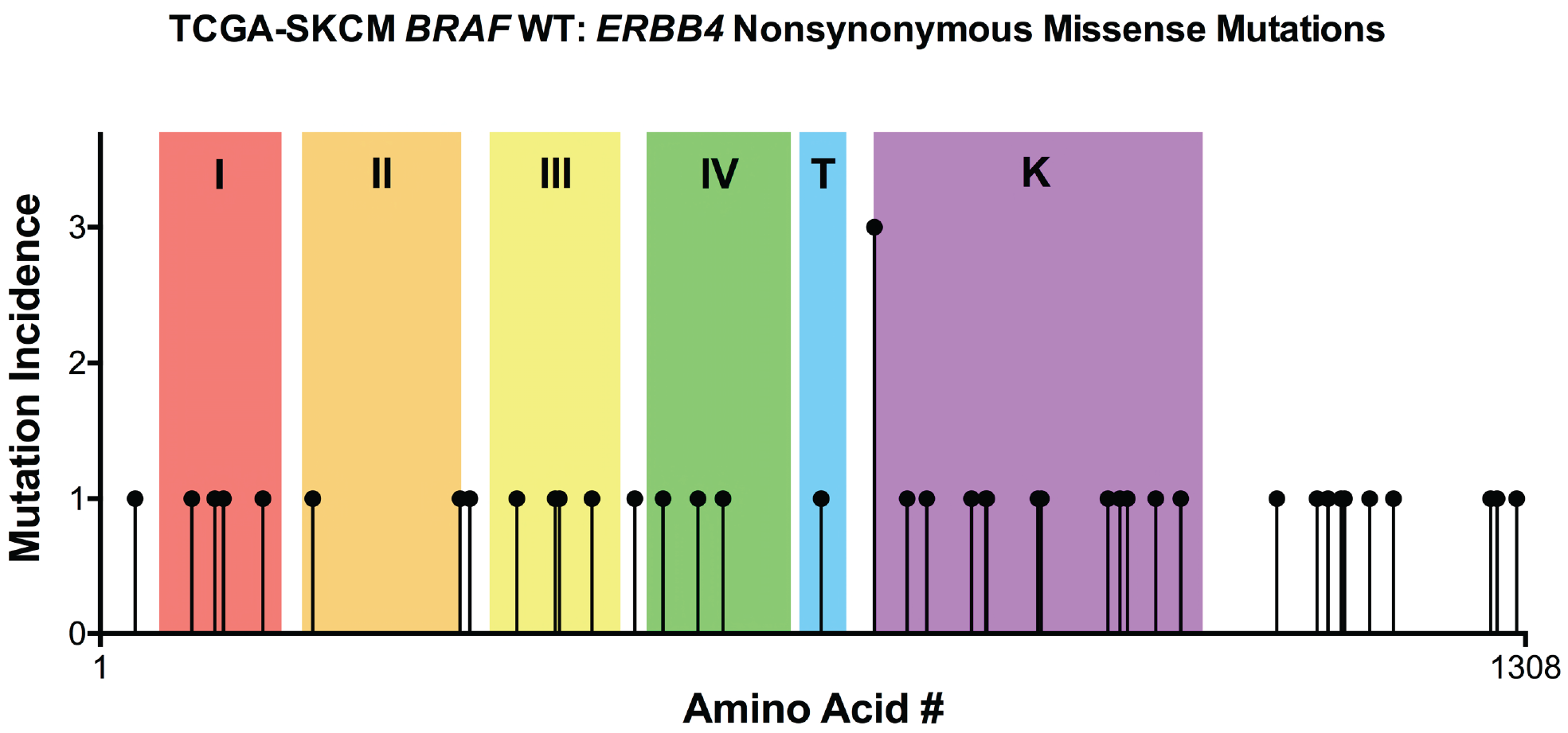
In *BRAF* WT melanomas of the TCGA-SKCM dataset, *ERBB4* nonsynonymous missense mutant alleles are evenly distributed across the entire *ERBB4* coding region.

The conservation of a particular amino acid residue across functionally related proteins suggests that residue is critical for the shared function of the related proteins. Thus, we hypothesized that *ERBB4* melanoma driver mutations are more likely to affect conserved residues.

Many members of the epidermal growth factor family of peptide hormones bind to both ERBB3 and ERBB4 (**Figure 2**) [34]. Moreover, ERBB3 dimerization and ERBB4 dimerization are regulated by an identical mechanism (**Figure 1**) [9]. Cases found in the TCGA-SKCM *BRAF* WT melanoma dataset possess fifteen different *ERBB4* missense mutations that alter amino acid residues found in the ERBB4 extracellular subdomains (ECDs) I-IV. We hypothesized that these fifteen residues correlated with ERBB4 residues that are conserved in the ERBB3 ECDs I-IV. However, only eleven residues altered by *ERBB4 BRAF* WT melanoma missense mutations are conserved in the ERBB3 ECDs I-IV, which falls well short of a statistically significant correlation (p=0.3763) (**Table 2a**). Nonetheless, these eleven *ERBB4* missense mutant alleles (G85S, R106C, R196C, P331S, G340E, A383T, S418F, T422I, E452K, R491K, and P517A) are assigned a priority point to help identify the best candidates for *ERBB4* driver mutant alleles in *BRAF* WT melanomas.

**Table 2a.**
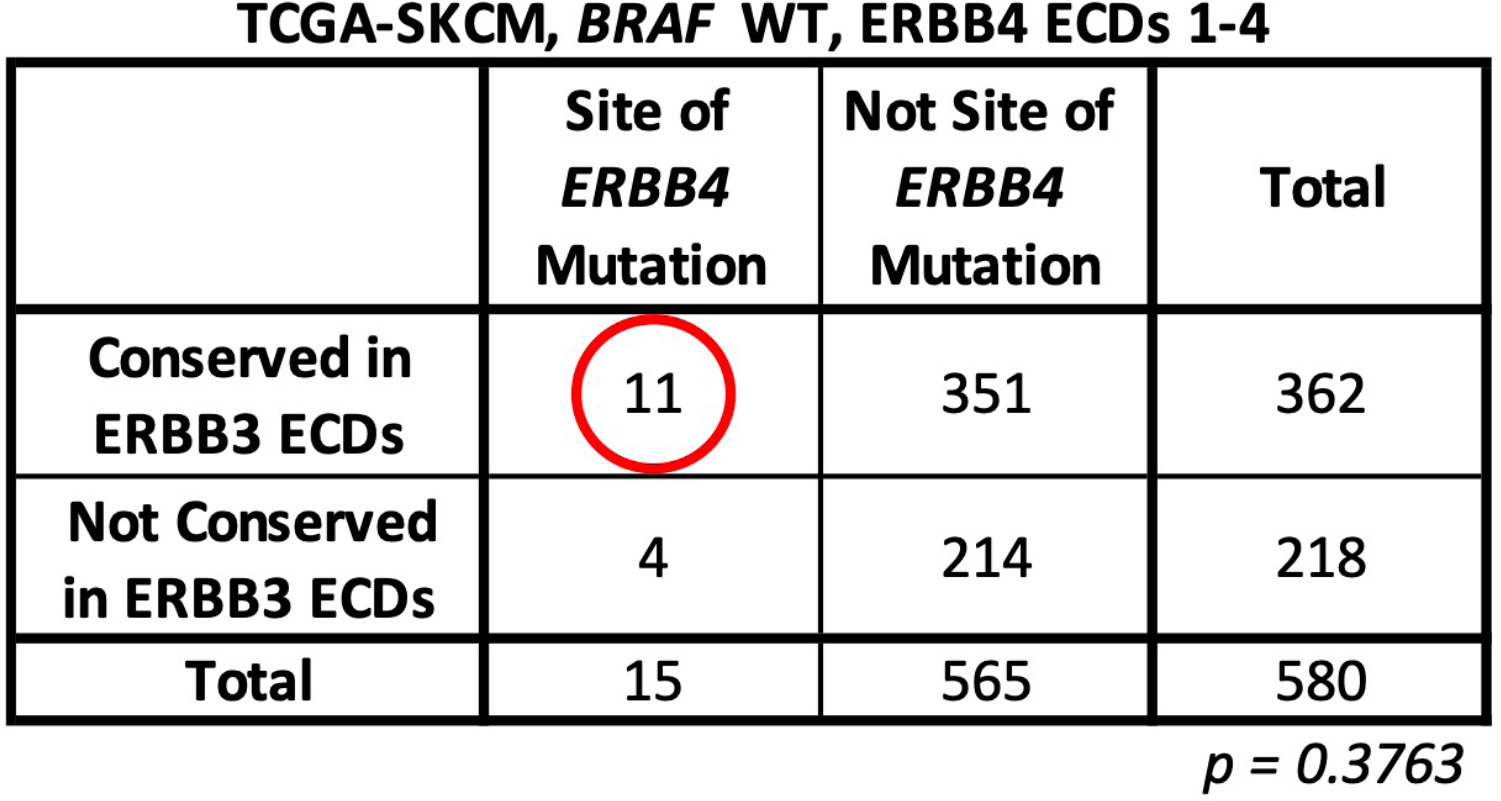
*ERBB4* missense mutations that affect residues of the extracellular domains (ECDs) are slightly correlated with residues that are conserved in the ERBB3 extracellular domains.

Only ERBB4 and EGFR directly bind ligands and form homodimers that undergo ligand-dependent receptor cross-phosphorylation [34]. (ERBB2 does not bind ligand and ERBB3 lacks sufficient kinase activity.) Cases found in the TCGA-SKCM *BRAF* WT melanoma dataset possess thirteen different *ERBB4* missense mutations that alter amino acid residues found in the ERBB4 tyrosine kinase domain. We hypothesized that these thirteen residues correlated with ERBB4 residues that are conserved in the EGFR tyrosine kinase domain. Indeed, all thirteen residues affected by *ERBB4 BRAF* WT melanoma missense mutations are conserved in the EGFR tyrosine kinase domain. However, this distribution falls just short (p=0.0565) of a statistically significant correlation (**Table 2b**). Nonetheless, these thirteen *ERBB4* missense mutant alleles (R711C, G741E, P759L, P800L, D813N, N814T, D861N, L864P, P925S, G936E, P943S, E969K, and R992C) are assigned a priority point to help identify the best candidates for *ERBB4* driver mutant alleles in *BRAF* WT melanomas.

**Table 2b.** *ERBB4* missense mutations that affect residues of the kinase domain are somewhat correlated with residues that that are conserved in the EGFR kinase domain.

**TCGA-SKCM, BRAF WT, ERBB4 Kinase Domain**
|  | Site of<br>*ERBB4*<br>Mutation | Not Site of<br>*ERBB4*<br>Mutation | Total |
| --- | --- | --- | --- |
| Conserved in<br>EGFR kinase<br>domain | 13 | 226 | 239 |
| Not Conserved<br>in EGFR kinase<br>domain | 0 | 64 | 64 |
| **Total** | **13** | **290** | **303** |

$p = 0.0565$

Finally, three *ERBB4* missense mutant alleles (R106C, G741E, L864P) found in the TCGA-SKCM *BRAF* WT melanoma data set correspond to gain-of-function alleles in *EGFR* (R108K, G735S, and L858R, respectively) or *ERBB2* (L866M corresponds to *ERBB4* L864P). As a result, these three *ERBB4* missense alleles (**Table 3**) are assigned a priority point to help identify the best candidates for *ERBB4* driver mutant alleles in *BRAF* WT melanomas.

**Table 3.** Three *ERBB4* missense mutations occur at residues that correspond to sites of oncogenic mutations in *EGFR* or *ERBB2*.

| *ERBB4* Mutation | Corresponding Mutation in *EGFR* | Corresponding Mutation in *ERBB2* | Function |
| --- | --- | --- | --- |
| R106C | R108K |  | Gain-of-function<br>Oncogenic |
| G741E | G735S |  | Gain-of-function<br>Oncogenic |
| L864P | L858R | L866M | Gain-of-function<br>Oncogenic |

### C. ERBB4 missense mutants are somewhat less common in TCGA-SKCM BRAF WT melanoma cases in which there are PI3K or PTEN alterations that are predicted to stimulate the PI3K pathway

The PI3K pathway is required for ERBB4-EGFR heterodimers to stimulate IL3-independent proliferation of BaF3 mouse lymphoid cells [35]. Likewise, an *ERBB4* gain-of-function mutant allele causes increased AKT phosphorylation in a human melanoma cell line [30]. Thus, we postulated that *ERBB4* driver mutants in BRAF WT melanomas stimulate PI3K pathway signaling and are inversely correlated with events (increased *PIK3CA* transcription, gain-of-function mutation in *PIK3CA*, decreased *PTEN* transcription, or loss-of-function mutation in *PTEN*) that would cause ERBB4-independent increases in PI3K pathway signaling. Twenty-nine cases in the TCGA-SKCM *BRAF* WT melanoma dataset contain an *ERBB4* missense mutation, but no *PIK3CA* mutation or increase in *PIK3CA* transcription, nor any *PTEN* mutation or decrease in *PTEN* transcription. However, this distribution falls just short (p=0.068) of a statistically significant correlation (**Table 4**).

**Table 4.** *ERBB4* missense mutants are somewhat inversely correlated with events (mutation of *PIK3CA*, increased *PIK3CA* transcription, mutation of *PTEN*, or decrease in *PTEN* transcription) predicted to result in increased PI3K pathway signaling activity.

|  | *ERBB4* Missense Mutation | *ERBB4* WT | Total |
| --- | --- | --- | --- |
| Mutation or Increase in *PIK3CA* Expression or Mutation or Decrease in *PTEN* Expression | 4 | 51 | 55 |
| No Mutation or Increase in *PIK3CA* Expression and <u>no</u> Mutation or Decrease in *PTEN* Expression | 29 | 138 | 167 |
| Total | 33 | 189 | 222 |

$p = 0.068$

### D. ERBB4 missense mutants are significantly more common in TCGA-SKCM BRAF WT melanoma cases in which there are RAS or NF1 nonsynonymous mutations

Gain-of-function *RAS* gene mutations occur in about 30% of skin cutaneous melanomas, and loss-of-function mutations in *NF1* occur in about 20% of skin cutaneous melanomas. Moreover, gain-of-function *BRAF* mutations, gain-of-function *RAS* gene mutations, and loss-of-function *NF1* mutations are largely mutually exclusive in skin cutaneous melanomas [36].

Receptor tyrosine kinases typically stimulate RAS pathway signaling [37–43]. Hence, in the *BRAF* WT melanomas of the TCGA-SKCM dataset, we predicted that cases that contain an ERBB4 missense mutation would be less likely to contain a nonsynonymous mutation in a *RAS* gene or *NF1*. Surprisingly, 29 cases in the TCGA-SKCM *BRAF* WT melanoma dataset contain an *ERBB4* missense mutation, as well as a nonsynonymous mutation in a *RAS* gene or *NF1* (Table 5). This statistically significant correlation (p=0.0189) suggests that ERBB4 signaling cooperates with RAS signaling to drive *BRAF* WT melanomas. Hence, we have assigned a priority point to the 33 *ERBB4* mutant alleles (E33K, G85S, R106C, D150N, P331S, A383T, S418F, T422I, E452K, R491K, P517A, G549S, P572L, F662L, R711C, P759L, P800L, D813N, D861N, P925S, G936E, E969K, R992C, P1080L, P1117L, R1127K, R1139Q, R1142Q, P1165L, E1187D, P1276S, P1282S, and P1300S) found in the 29 cases that contain a nonsynonymous mutation in a *RAS* gene or *NF1*.

**Table 5.** *ERBB4* missense mutations are significantly correlated with a *RAS* or *NF1* nonsynonymous mutation.

|  | *ERBB4* Missense Mutation | *ERBB4* WT | Total |
| --- | --- | --- | --- |
| *RAS* or *NF1* Nonsynonymous Mutation | 29 | 128 | 157 |
| *RAS* and *NF1* WT | 4 | 61 | 65 |
| Total | 33 | 189 | 222 |

$p = 0.0189$

### E. ERBB4 missense mutations are significantly more likely in cases where there is a RAS or NF1 nonsynonymous mutation as well as no other apparent cause of increased PI3K signaling

Our *in silico* analyses lead us to hypothesize that gain-of-function *ERBB4* mutants stimulate signaling by the PI3K pathway and that this PI3K signaling cooperates with elevated RAS signaling (caused by a gain-of-function *RAS* gene mutation or a loss-of-function *NF1* mutation) to drive *BRAF* WT melanomas. Thus, we tested whether *ERBB4* missense mutations are more likely to occur in cases where there is a *RAS* gene or *NF1* mutation and NO events (increases in *PIK3CA* transcription, gain-of-function mutations in *PIK3CA*, decreases in *PTEN* transcription, or loss-of-function mutations in *PTEN*) that would cause ERBB4-independent increases in PI3K pathway signaling (**Table 6**). Twenty-five cases in the TCGA-SKCM *BRAF* WT melanoma dataset contain an *ERBB4* missense mutation, as well as a nonsynonymous mutation in a *RAS* gene or *NF1*, but no event (increases in *PIK3CA* transcription, gain-of-function mutations in *PIK3CA*, decreases in *PTEN* transcription, or loss-of-function mutations in *PTEN*) that would cause ERBB4-independent increases in PI3K pathway signaling. This statistically significant correlation (p=0.0166) supports our hypothesis. Thus, the 29 *ERBB4* mutant alleles (E33K, G85S, R106C, D150N, P331S, A383T, S418F, T422I, E452K, R491K, P517A, G549S, R711C, P759L, P800L, D813N, D861N, P925S, G936E, E969K, R992C, P1080L, R1127K, R1139Q, P1165L, E1187D, P1276S, P1282S, and P1300S) found in these 25 cases are assigned a priority point to help identify the best candidates for *ERBB4* driver mutant alleles in *BRAF* WT melanomas.

**Table 6.** ERBB4 missense mutations are significantly correlated with a *RAS* or *NF1* mutation AND no other apparent cause of increased PI3K activity.

|  |  | *ERBB4*<br>Missense<br>Mutation | *ERBB4* WT | Totals |
| --- | --- | --- | --- | --- |
| *RAS* or *NF1*<br>Mutation | *PIK3CA* Mutation or Increase<br>in *PIK3CA* Expression or *PTEN* Mutation or<br>Decrease in *PTEN* Expression | 4 | 35 | 39 |
|  | No *PIK3CA* Mutation or Increase<br>in *PIK3CA* Expression and no *PTEN*<br>Mutation or Decrease in *PTEN* Expression | 25 | 93 | 118 |
| *RAS* and *NF1*<br>WT | No Consideration of *PIK3CA* /*PTEN*<br>Mutation/Expression Status | 4 | 61 | 65 |
| Totals |  | 33 | 189 | 222 |

$p = 0.016$

### F. ERBB4 mutations and elevated ERBB4 transcription appear to independently drive BRAF WT melanomas

*In silico* analyses of the TCGA-SKCM *BRAF* WT melanoma dataset suggest that elevated *ERBB4* transcription appears to cooperate with elevated RAS signaling (due to a gain-of-function *RAS* gene mutation or a loss-of-function *NF1* mutation) to drive *BRAF* WT melanomas [8]. Thus, it is possible that *ERBB4* mutant alleles require elevated *ERBB4* transcription to drive *BRAF* WT melanomas. We have identified 129 cases in the TCGA-SKCM *BRAF* WT melanoma dataset that contain a non-synonymous *RAS* or *NF1* mutation and for which *ERBB4* transcription data is available. Seventeen of these cases contain an *ERBB4* missense mutation but do not exhibit elevated *ERBB4* transcription; nineteen cases do not contain an *ERBB4* missense mutation but do exhibit elevated *ERBB4* transcription; and three cases contain an *ERBB4* missense mutation and exhibit elevated *ERBB4* transcription (**Figure 4**). These data suggest that *ERBB4* mutations and elevated *ERBB4* transcription appear to independently drive *BRAF* WT melanomas. Seventeen *ERBB4* mutant alleles were found in the seventeen cases that contain an *ERBB4* missense mutation but do not exhibit elevated *ERBB4* transcription. These seventeen *ERBB4* mutant alleles (R106C, D150N, A383T, S418F, R491K, P517A, G549S, F662L, R711C, P759L, P800L, D861N, G936E, E969K, R1142Q, P1276S, P1300S) are assigned a priority point to help identify the best candidates for *ERBB4* driver mutant alleles in *BRAF* WT melanomas.

**Figure 4.**
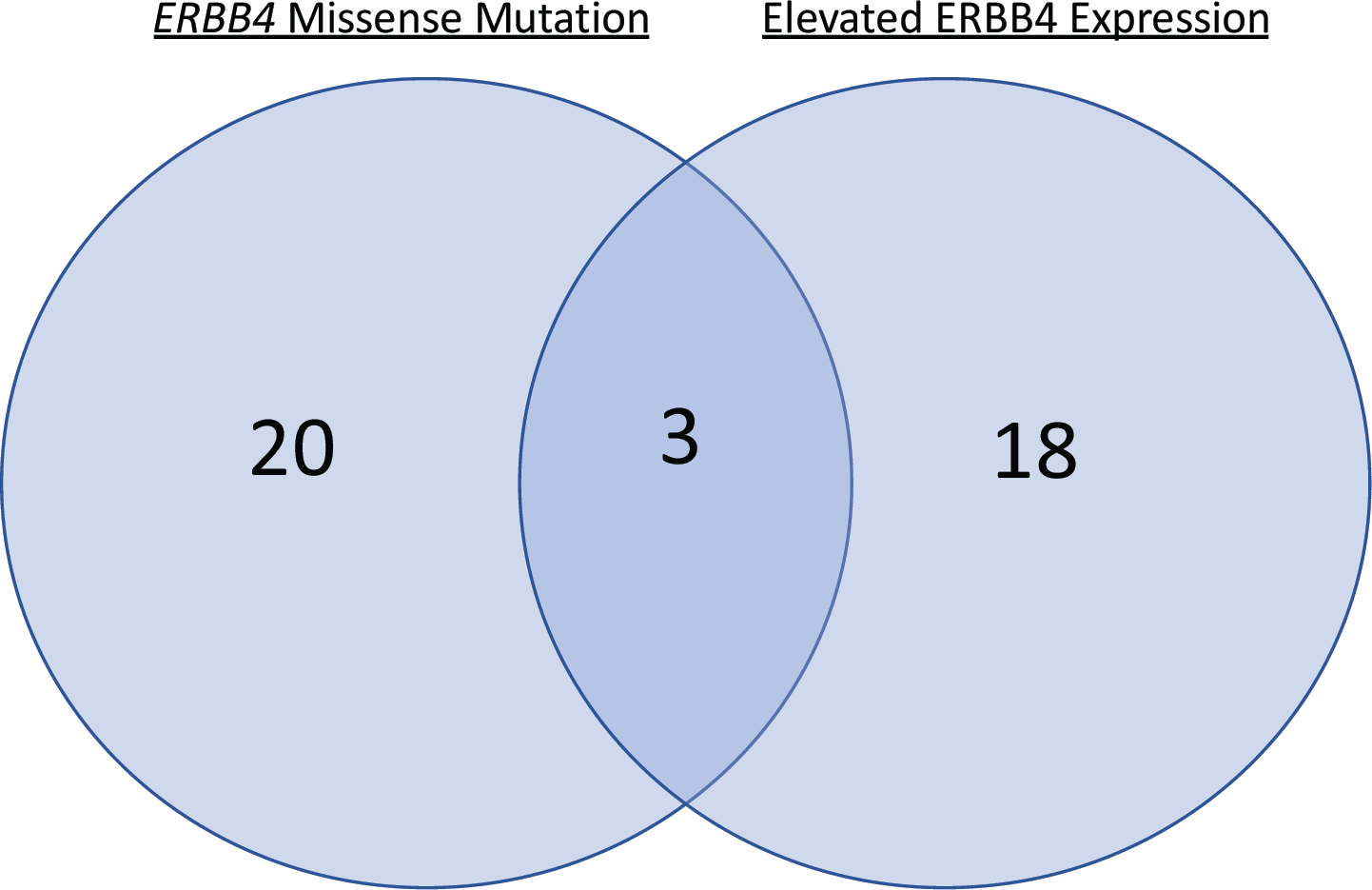
Within the 129 *BRAF* WT cases in the TCGA-SKCM dataset that harbor a *RAS* or *NF1* nonsynonymous mutation, an *ERBB4* missense mutation and elevated *ERBB4* transcription appear to be largely mutually exclusive.

**Figure 5.**
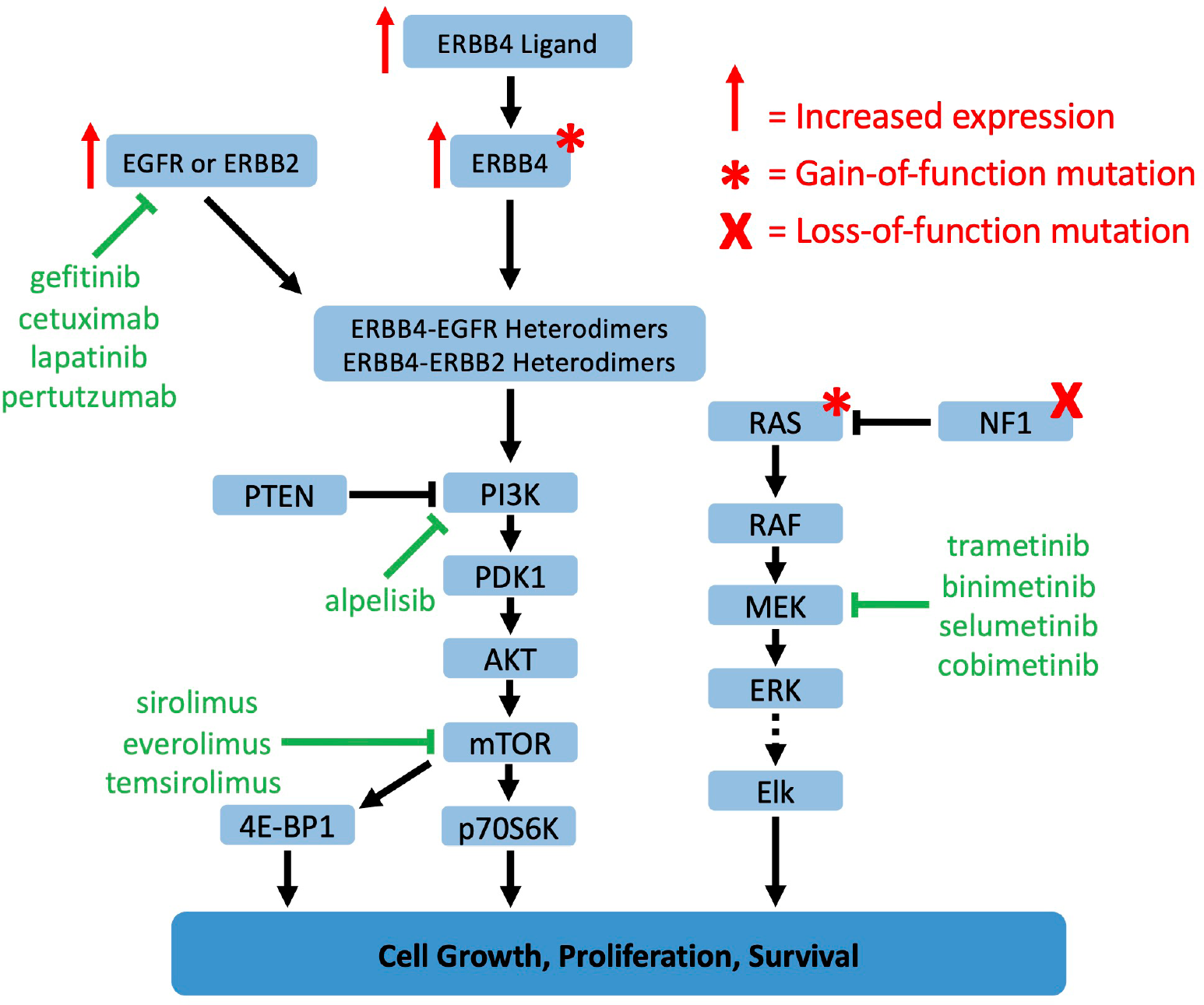
We predict that gain-of-function *ERBB4* mutations in *BRAF* WT tumors cause elevated signaling by ERBB4-ERBB2 or ERBB4-EGFR heterodimers, which stimulates signaling by the PI3K pathway, which cooperates with elevated RAS signaling to drive *BRAF* WT tumors. Adapted from [8].

### G. Among deceased BRAF WT melanoma patients of the TCGA-SKCM dataset, ERBB4 mutant alleles appear to be associated with a gender disparity

Our previous work indicated that males are more likely to die of BRAF WT melanomas than females [8]. Here we explored the role that *ERBB4* mutations may play in that gender disparity. In the *BRAF* WT cases of the TCGA-SKCM dataset, *ERBB4* mutational status is available for 115 deceased patients (79 male and 36 female – **Table 7**). Of the 36 deceased female *BRAF* WT melanoma patients, only one possessed a melanoma that harbored an *ERBB4* mutant allele. In contrast, of the 79 deceased male *BRAF* WT melanoma patients, 15 possessed a melanoma that harbored an *ERBB4* mutant allele. This association of an *ERBB4* mutation with males among deceased *BRAF* WT melanoma patients is statistically significant and suggests elevated ERBB4 signaling may play a greater role among male *BRAF* WT melanoma patients than female *BRAF* WT melanoma patients. Fifteen different *ERBB4* mutant alleles were found in the 15 cases of deceased male *BRAF* WT melanoma patients who possessed an *ERBB4* mutation. These fifteen *ERBB4* mutant alleles (D150N, A383T, T422I, E452K, F662L, P759L, P800L, N814T, P925S, P1117L, R1142Q, E1187D, P1282S, P1276S) are assigned a priority point to help identify the best candidates for *ERBB4* driver mutant alleles in *BRAF* WT melanomas.

**Table 7.**
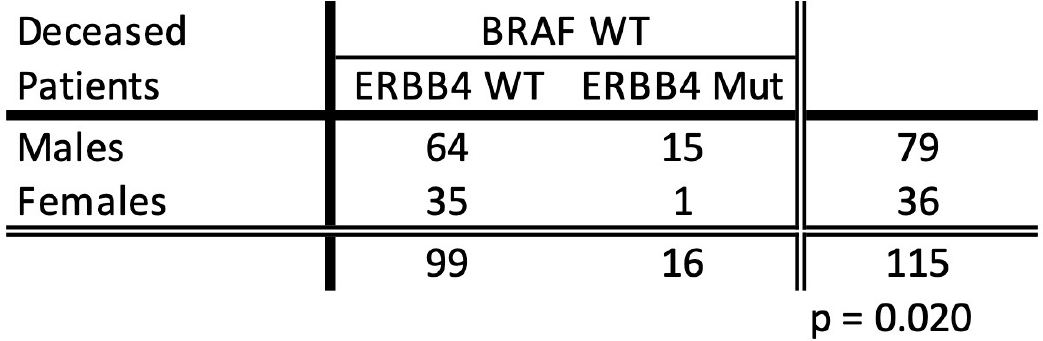
Among deceased *BRAF* WT melanoma patients, an *ERBB4* mutation is associated with male patients to a much greater extent than female patients.

| Deceased<br>Patients | BRAF WT |  |  |
| --- | --- | --- | --- |
|  | *ERBB4* WT | *ERBB4* Mut |  |
| Males | 64 | 15 | 79 |
| Females | 35 | 1 | 36 |
|  | 99 | 16 | 115 |

$p = 0.020$

### H. We have prioritized ERBB4 mutant alleles as candidate drivers of BRAF WT melanomas

Based on the *in silico* data presented in **Table 2a, Table 2b, Table 3, Table 5, Table 6, Figure 4**, and **Table 7**, we have prioritized *ERBB4* mutant alleles as candidate drivers of *BRAF* WT melanomas (**Table 8**). Next, we will describe our efforts to determine whether selected *ERBB4* mutant alleles (shown in green in **Table 8**) function as drivers of *BRAF* WT melanomas. Doing so will evaluate the accuracy of our prioritization scheme in predicting which *ERBB4* mutant alleles function as *bona fide BRAF* WT melanoma tumor drivers.

**Table 8.**
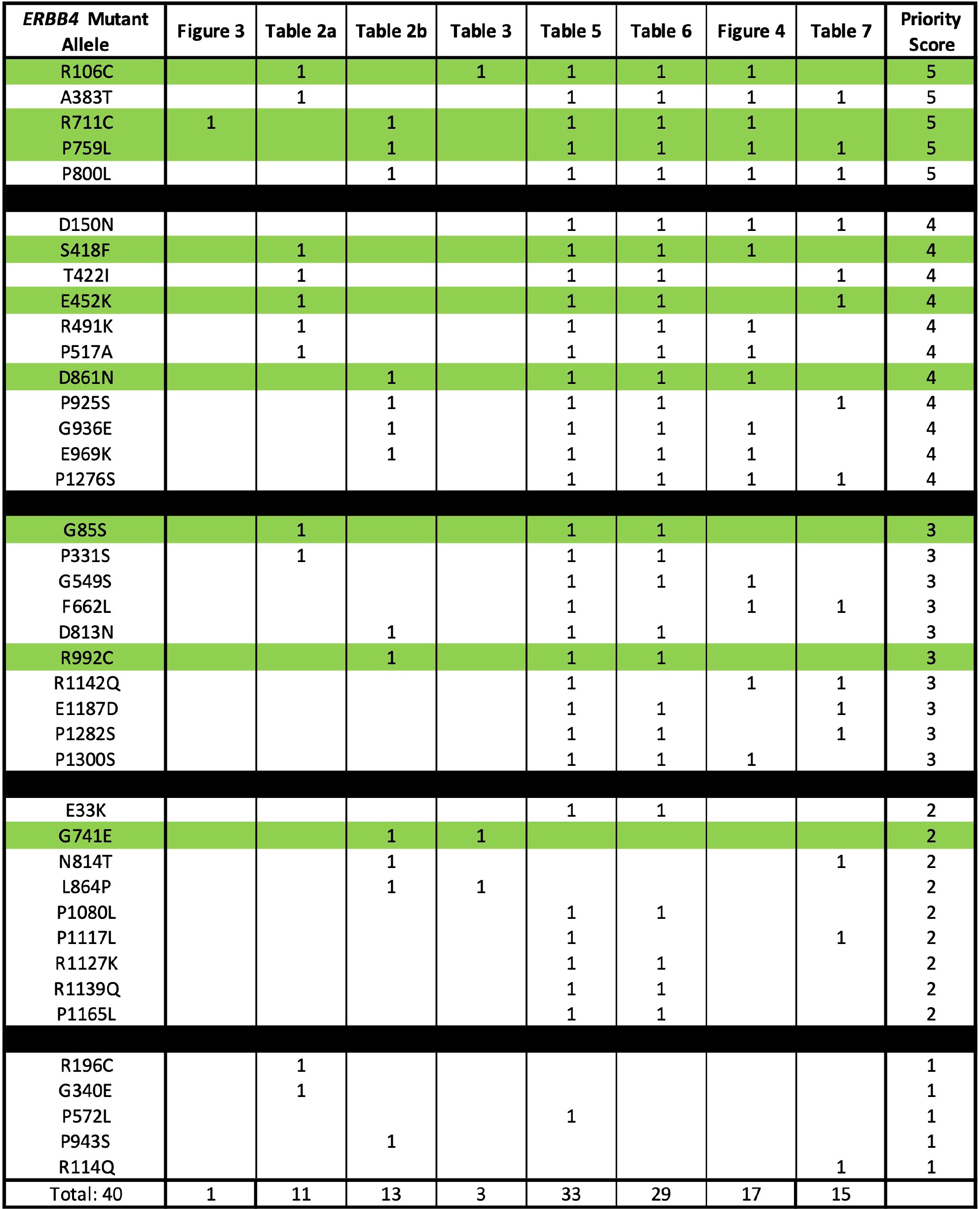
Prioritization of candidate *ERBB4* driver mutant alleles in the TCGA-SKCM *BRAF* WT melanoma dataset. These data are extracted from the tables and figures (and associated text) indicated in the table. Green highlighting indicates a mutant allele that has been tested for driver activity.

| *ERBB4* Mutant Allele | Figure 3 | Table 2a | Table 2b | Table 3 | Table 5 | Table 6 | Figure 4 | Table 7 | Priority Score |
| --- | --- | --- | --- | --- | --- | --- | --- | --- | --- |
| R106C |  | 1 |  | 1 | 1 | 1 | 1 |  | 5 |
| A383T |  | 1 |  |  | 1 | 1 | 1 | 1 | 5 |
| R711C | 1 |  | 1 |  | 1 | 1 | 1 |  | 5 |
| P759L |  |  | 1 |  | 1 | 1 | 1 | 1 | 5 |
| P800L |  |  | 1 |  | 1 | 1 | 1 | 1 | 5 |
| D150N |  |  |  |  | 1 | 1 | 1 | 1 | 4 |
| S418F |  | 1 |  |  | 1 | 1 | 1 |  | 4 |
| T422I |  | 1 |  |  | 1 | 1 |  | 1 | 4 |
| E452K |  | 1 |  |  | 1 | 1 |  | 1 | 4 |
| R491K |  | 1 |  |  | 1 | 1 | 1 |  | 4 |
| P517A |  | 1 |  |  | 1 | 1 | 1 |  | 4 |
| D861N |  |  | 1 |  | 1 | 1 | 1 |  | 4 |
| P925S |  |  | 1 |  | 1 | 1 |  | 1 | 4 |
| G936E |  |  | 1 |  | 1 | 1 | 1 |  | 4 |
| E969K |  |  | 1 |  | 1 | 1 | 1 |  | 4 |
| P1276S |  |  |  |  | 1 | 1 | 1 | 1 | 4 |
| G85S |  | 1 |  |  | 1 | 1 |  |  | 3 |
| P331S |  | 1 |  |  | 1 | 1 |  |  | 3 |
| G549S |  |  |  |  | 1 | 1 | 1 |  | 3 |
| F662L |  |  |  |  | 1 |  | 1 | 1 | 3 |
| D813N |  |  | 1 |  | 1 | 1 |  |  | 3 |
| R992C |  |  | 1 |  | 1 | 1 |  |  | 3 |
| R1142Q |  |  |  |  | 1 |  | 1 | 1 | 3 |
| E1187D |  |  |  |  | 1 | 1 |  | 1 | 3 |
| P1282S |  |  |  |  | 1 | 1 |  | 1 | 3 |
| P1300S |  |  |  |  | 1 | 1 | 1 |  | 3 |
| E33K |  |  |  |  | 1 | 1 |  |  | 2 |
| G741E |  |  | 1 | 1 |  |  |  |  | 2 |
| N814T |  |  | 1 |  |  |  |  | 1 | 2 |
| L864P |  |  | 1 | 1 |  |  |  |  | 2 |
| P1080L |  |  |  |  | 1 | 1 |  |  | 2 |
| P1117L |  |  |  |  | 1 |  |  | 1 | 2 |
| R1127K |  |  |  |  | 1 | 1 |  |  | 2 |
| R1139Q |  |  |  |  | 1 | 1 |  |  | 2 |
| P1165L |  |  |  |  | 1 | 1 |  |  | 2 |
| R196C |  | 1 |  |  |  |  |  |  | 1 |
| G340E |  | 1 |  |  |  |  |  |  | 1 |
| P572L |  |  |  |  | 1 |  |  |  | 1 |
| P943S |  |  | 1 |  |  |  |  |  | 1 |
| R114Q |  |  |  |  |  |  |  | 1 | 1 |
| Total: 40 | 1 | 11 | 13 | 3 | 33 | 29 | 17 | 15 |  |

### I. Two ERBB4 mutant alleles (G85S and G741E) found in BRAF WT melanoma samples cause increased clonogenic proliferation of the MEL-JUSO BRAF WT melanoma cell line

We have shown that ectopic expression of wild-type *ERBB4* (*ERBB4* WT) significantly stimulates the clonogenic proliferation of IPC-298, MEL-JUSO, MeWo, and SK-MEL-2 human *BRAF* WT melanoma cells [8]. Moreover, ectopic expression of the dominant-negative (DN) *ERBB4* K751M mutant allele significantly inhibits the clonogenic proliferation of these same cells [8]. These results indicate that *ERBB4* is both sufficient and necessary to drive some *BRAF* WT melanoma cell lines; they also suggest that we can use the IPC-298, MEL-JUSO, MeWo, and SK-MEL-2 cell lines to identify *ERBB4* mutant alleles that function as *bona fide* drivers of *BRAF* WT melanomas.

We have evaluated the ability of selected *ERBB4* mutant alleles found in *BRAF* WT melanomas to stimulate clonogenic proliferation of the MEL-JUSO cell line (**Table 9**). One positive control is *ERBB4* WT, which stimulates clonogenic proliferation of MEL-JUSO cells relative to the vector control. Another positive control is the *ERBB4* Y285C mutant allele, which has been found in non-small cell lung carcinoma samples and stimulates ligand-dependent and -independent ERBB4 signaling and increased ERBB4 heterodimerization with ERBB2 [9, 44]. A third control is the *ERBB4* Q646C mutant allele, which is constitutively homodimerized and tyrosine phosphorylated and appears to function as a tumor suppressor; it inhibits the proliferation of many cell lines derived from solid tumors [45–50]. These controls performed as anticipated (**Table 9**). *ERBB4* WT significantly stimulates clonogenic proliferation of the MEL-JUSO cell line relative to the vector control. Likewise, the *ERBB4* Y285C mutant allele significantly stimulates clonogenic proliferation of the MEL-JUSO cell line relative to *ERBB4* WT. And, relative to the vector control, the *ERBB4* Q646C mutant allele significantly inhibits clonogenic proliferation of the MEL-JUSO cell line.

**Table 9.** The *ERBB4* G85S and G741E mutant alleles found in *BRAF* WT melanomas drive increased clonogenic proliferation of the MEL-JUSO *BRAF* WT melanoma cell line.

| Construct | Average Efficiency of Clonogenic Proliferation Relative to *ERBB4* WT | p of 1-Tailed, 1-Sample t-test Compared to *ERBB4* WT | N |
| --- | --- | --- | --- |
| Vector Control | 36% | 3.34E-04 | 4 |
| *ERBB4* WT | 100% |  | 12 |
| *ERBB4* G85S | 166% | 0.001 | 11 |
| *ERBB4* R106C | 95% | 0.248 | 12 |
| *ERBB4* Y285C | 156% | 0.006 | 10 |
| *ERBB4* S418F | 92% | 0.137 | 11 |
| *ERBB4* E452K | 100% | 0.489 | 11 |
| *ERBB4* Q646C* (Relative to Vector) | 29% | 0.008 | 3 |
| *ERBB4* R711C | 166% | 0.056 | 10 |
| *ERBB4* G741E | 148% | 0.008 | 11 |
| *ERBB4* P759L | 115% | 0.232 | 12 |
| *ERBB4* D861N | 114% | 0.089 | 10 |
| *ERBB4* R992C | 112% | 0.188 | 11 |

The *ERBB4* G85S and G741E mutant alleles significantly stimulate clonogenic proliferation of the MEL-JUSO cell line relative to *ERBB*4 WT (**Table 9**). Thus, the *ERBB4* G85S and G741E mutant alleles appear to function as *bona fide* drivers of some *BRAF* WT melanomas. However, the other seven *ERBB4* mutant alleles found in *BRAF* WT melanoma samples (R106C, S418F, E452K, R711C, P759L, D861N, and R992C) failed to significantly stimulate clonogenic proliferation, although R711C and D861N almost caused a significant effect.

Earlier in this paper we noted that *ERBB4* mutant alleles may play a greater role in *BRAF* WT melanomas in males than in females. The IPC-298 and MEL-JUSO cell lines were generated from tumors of female *BRAF* WT melanoma patients, whereas the MeWo and SK-MEL-2 cell lines were generated from tumors of male *BRAF* WT melanoma patients. Hence, the MeWo and SK-MEL-2 cell lines may be more appropriate for evaluating the biological activities of the *ERBB4* mutant alleles.

Thus, we are evaluating whether *ERBB4* mutant alleles found in BRAF WT melanomas stimulate the clonogenic proliferation of MeWo and SK-MEL-2 cell lines (**Table 10**). Thus far, the activities of the control *ERBB4* alleles (WT, Y285C, and Q646C) in the MeWo and SK-MEL-2 cell lines are qualitatively identical to the activities of these alleles in the MEL-JUSO cell line. Likewise, it appears that the *ERBB4* G85S and G741E alleles, which exhibit a gain-of-function phenotype in the MEL-JUSO cell line, also exhibit a gain-of-function phenotype in the MeWo and SK-MEL-2 cell line. Thus, for now it appears that the MEL-JUSO, MeWo, and SK-MEL-2 cells respond in a qualitatively similar manner to the various *ERBB4* mutant alleles. However, several more trials are needed to properly evaluate the activities of the *ERBB4* mutant alleles in the MeWo and SK-MEL-2 cell lines.

**Table 10.**
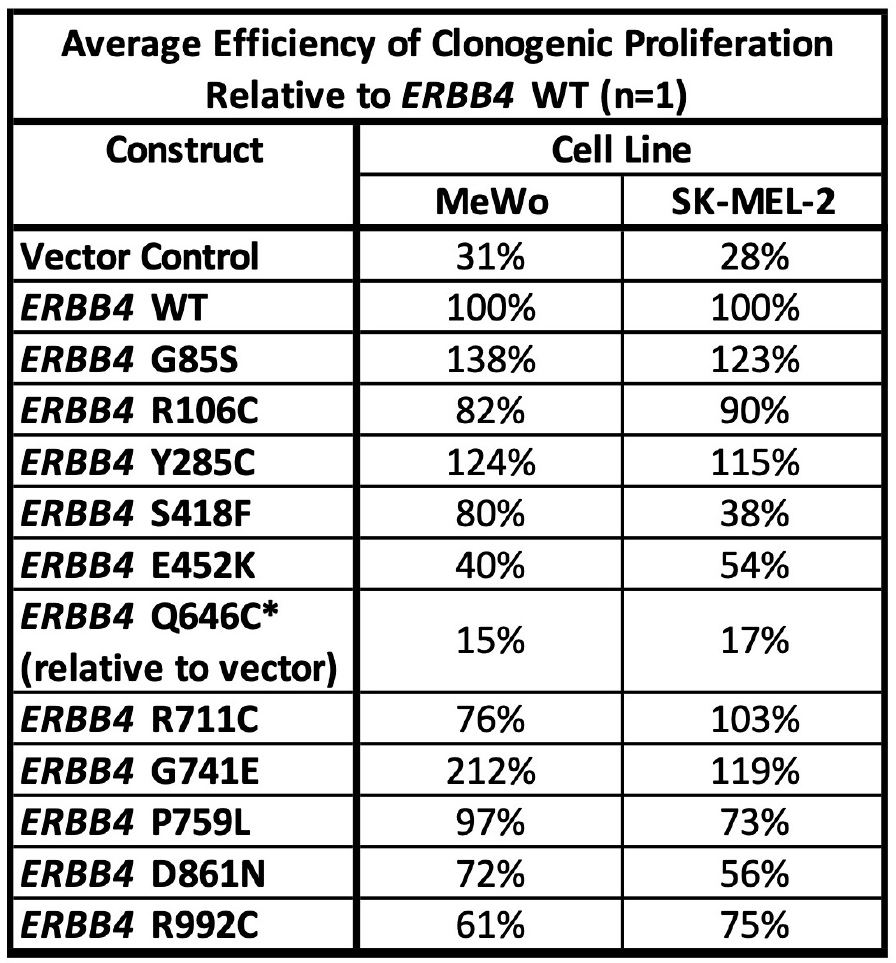
Preliminary data suggest that the *ERBB4* G85S and G741E mutant alleles found in *BRAF* WT melanomas drive increased clonogenic proliferation of the MeWo and SK-MEL-2 female *BRAF* WT melanoma cell lines.

### J. A heterologous model system can be used to determine whether ERBB4 mutants found in BRAF WT melanoma samples exhibit elevated ligand-dependent or ligand-independent ERBB4 signaling activity

The IPC-298, MEL-JUSO, MeWo, and SK-MEL-2 cell lines exhibit endogenous expression of ERBB4 ligands [8]. Thus, *ERBB4* mutants could stimulate clonogenic proliferation of these cell lines via a ligand-dependent or ligand-independent mechanism.

We and others have previously demonstrated that ERBB4 homodimers typically do not stimulate cell proliferation, whereas ERBB4-ERBB2 and ERBB4-EGFR heterodimers do stimulate cell proliferation [9]. Indeed, ERBB4 ligands do not stimulate proliferation of BaF3 mouse lymphoid cells engineered to express ERBB4 but do stimulate proliferation of BaF3 cells engineered to express ERBB4 and EGFR or ERBB4 and ERBB2 [9, 51]. BaF3 cells lack endogenous EGFR, ERBB2, and ERBB4 expression. But, because they endogenously express ERBB3, they are not ideal for studying the functional differences among ERBB4 homodimers, ERBB4-EGFR heterodimers, or ERBB4-ERBB2 heterodimers [51].

Thus, we have chosen to study these functional differences using derivatives of the 32D mouse lymphoid cell line, which lack endogenous expression of all four ERBB family receptors and are dependent on interleukin 3 (IL3) for their proliferation [52]. We have demonstrated that ERBB2, but not EGFR, is required for clonogenic proliferation of the MEL-JUSO BRAF WT melanoma cell line [8]. Thus, 32D cell lines were generated that express both *ERBB4* and *ERBB2*, *ERBB4* alone, *ERBB2* alone, and the appropriate vector controls. None of these 32D cell lines exhibit proliferation in the absence of both IL3 and an ERBB4 ligand. Moreover, the ERBB4 ligand NRG1beta stimulates IL3-independent proliferation of the 32D/ERBB2+ERBB4 cells but does not stimulate IL3-independent proliferation of any of the other 32D cell lines (**Table 11**). Thus, 32D cell lines that co-express ERBB4 mutant proteins and ERBB2 are ideal for studying whether the ERBB4 mutant proteins encoded by *ERBB4* mutant alleles found in *BRAF* WT melanomas potentiate the effects of ERBB4 ligands or enable ERBB4 coupling to proliferation independent of an ERBB4 ligand.

**Table 11.**
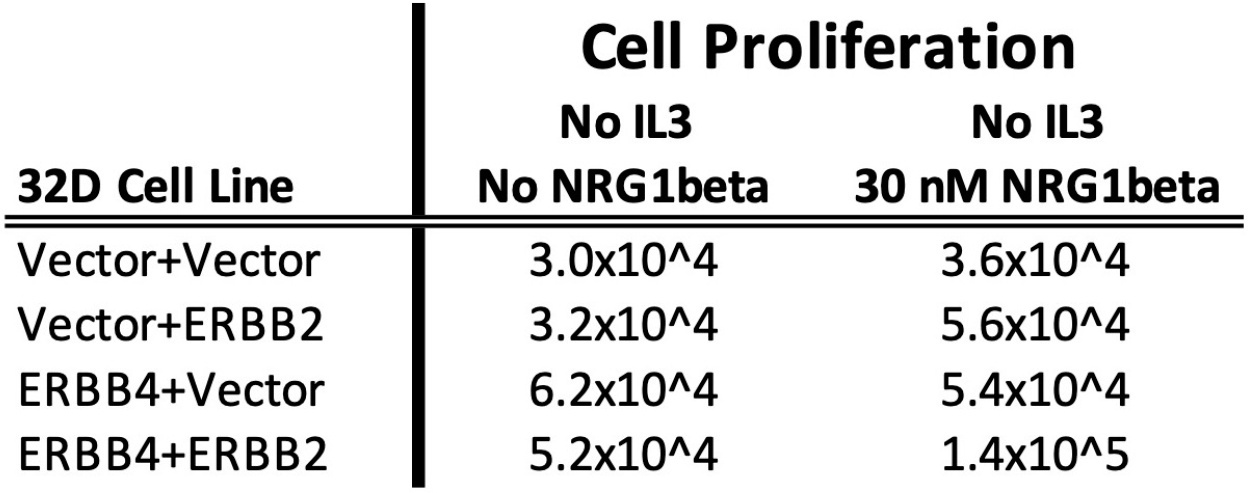
The ERBB4 ligand NRG1beta stimulates IL3-independent proliferation of the 32D/ERBB4+ERBB2 cell line, but not any of the control cell lines.

| 32D Cell Line | Cell Proliferation |  |
| --- | --- | --- |
|  | No IL3 | No IL3 |
|  | No NRG1beta | 30 nM NRG1beta |
| Vector+Vector | 3.0x10<sup>4</sup> | 3.6x10<sup>4</sup> |
| Vector+ERBB2 | 3.2x10<sup>4</sup> | 5.6x10<sup>4</sup> |
| ERBB4+Vector | 6.2x10<sup>4</sup> | 5.4x10<sup>4</sup> |
| ERBB4+ERBB2 | 5.2x10<sup>4</sup> | 1.4x10<sup>5</sup> |

## Materials and Methods

### Analysis of the TCGA-SKCM dataset

Clinical and biospecimen data were downloaded for all 470 cases found in the TCGA-SKCM dataset [31]. All analyzed datasets are publicly available through the NIH/NCI Genomic Data Commons (GDC) portal [53]. The R statistical computing and graphics environment software [54] was used to reorganize the dataset. Statistical analyses were performed using GraphPad Prism [55].

### Creating *BRAF* WT dataset

All *BRAF* WT cases were segregated from the TCGA-SKCM dataset for analysis. Cases in which there were ERBB4 mutation(s) which co-occurred with a stop gained or splice-site mutation were removed from the *BRAF* WT dataset for analysis of the effect of *ERBB4* missense mutation in *BRAF* WT melanoma (Sections IIIB-F). Three mutations fell into this category (M958I, R47Q, G573D). These mutations and the stop gained mutated cases are included in the analysis for section IIIA.

### Cell Lines and cell culture

Mouse C127 fibroblasts and the Ψ2 and PA317 recombinant retrovirus packaging cell lines are generous gifts of Daniel DiMaio (Yale University). These cells were cultured essentially as described previously [56]. The MEL-JUSO [57] human melanoma cell line was obtained from DSMZ [58] (Braunschweig, Germany) and was cultured as recommended. The 32D mouse myeloid cell line has been described previously and was cultured as recommended [59]. Cell culture media, serum, and supplements were obtained from Cytiva [60] (Marlborough, VA). G418 was obtained from Corning [61] (Corning, NY). Genetic and mRNA expression data for the MEL-JUSO cell line were obtained from the Broad Institute Cancer Cell Line Encyclopedia (CCLE) [62].

### Mutagenesis of DNA Constructs

The recombinant retroviral expression construct pLXSN-ERBB4 has been described previously [51]. We used pLXSN-ERBB4 as the parent plasmid for site-directed mutagenesis using the Q5 Site Directed Mutagenesis Kit (NEB). Mutagenesis was performed essentially as recommended by the manufacturer.

### Recombinant retroviruses

Briefly, the recombinant amphotropic retroviruses LXSN, LXSN-ERBB4 (ERBB4 WT), LXSN-ERBB4 R106C, LXSN-ERBB4 Y285C, and LXSN-ERBB4 P759L were packaged using the Ψ2 ecotropic retrovirus packaging cell line, and the PA317 amphotropic retrovirus packaging cell line as previously described [46, 51, 63]. 32D cells were infected with the recombinant amphotropic retroviruses LXSN, LXSN-ERBB4, LXSN-HygR, and LXSN-HygR-ERBB2 at a 0.1 multiplicity of infection. These constructs have been previously described [52, 56].

### Clonogenic Proliferation Assay

C127, MEL-JUSO cells were infected with 500 and 3000 amphotropic retroviral infectious units (respectively), essentially as described earlier. After incubation with the viruses, infected cells were selected using G418 at a concentration of 800 ug/mL. The resulting colonies of G418-resistant cells were stained using Giemsa 8 and 13 days later (respectively), and colonies were counted. C127 infections served as a control for viral titer. Tissue culture plates were digitized, and clonogenic proliferation efficiency was calculated as previously described [46]. The statistical significance of differences in clonogenic proliferation was calculated using ANOVA with a p-value threshold of <0.05 (1-tailed).

### Stimulation of ERBB4-ERBB2 heterodimer dependent proliferation in 32D cells

32D cells which stably express ERBB4, ERBB2, or ERBB4 and ERBB2 were mock treated or treated with 30 nM of NRG1β (Recombinant Human Heregulinβ-1, Peprotech) in the absence of IL3. After six days, cells were counted using a hemacytometer to evaluate NRG1β-dependent growth.

## Discussion

### A. The apparent selection for ERBB4 mutant alleles in the TCGA-SKCM BRAF WT melanoma data set enables prioritization of individual ERBB4 mutant alleles

Previous *in silico* analyses of the TCGA-SKCM BRAF WT melanoma dataset suggest that elevated *ERBB4* transcription drives the proliferation of *BRAF* WT melanomas [8]. In this work *in silico* analyses of *ERBB4* mutant alleles found in the TCGA-SKCM *BRAF* WT melanoma data set suggest that *ERBB4* mutant alleles may drive the proliferation of *BRAF* WT melanomas. Moreover, these analyses establish prioritization scheme (**Table 8**) for identifying those mutants that are most likely to function as BRAF WT melanoma drivers.

### B. The ERBB4 G85S and G741E mutant alleles found in BRAF WT melanoma samples stimulate clonogenic proliferation of the MEL-JUSO BRAF WT melanoma cell line

These results (**Table 9**) support our hypothesis that at least some *ERBB4* mutant alleles found in *BRAF* WT melanoma samples can drive *BRAF* WT melanoma cell lines. However, it is surprising that none of the higher-priority *ERBB4* mutant alleles (R106C, R711C, P759L, S418F, E452K, and D861N – **Table 8**) appear to significantly stimulate clonogenic proliferation of MEL-JUSO cells (**Table 9**). Because MEL-JUSO cells are derived from a female BRAF WT melanoma patient, we hypothesized that analyzing the activity of the ERBB4 mutant alleles in cell lines (MeWo, SK-MEL-2) derived from female BRAF WT melanoma patients might yield different results. However, preliminary results (**Table 10**) do not support that hypothesis.

### C. 32D/ERBB4+ERBB2 and control cell lines are suitable for determining which candidate ERBB4 gain-of-function mutants stimulate cell proliferation

It appears that our prioritization scheme (**Table 8**) may not accurately identify those *ERBB4* mutants found in *BRAF* WT melanomas that are most likely to function as drivers of these tumors. Likewise, the throughput of our clonogenic proliferation assays (**Table 9**) does not appear to be sufficient to effectively evaluate the 40 different *ERBB4* mutant alleles found in *BRAF* WT melanomas. Thus, we must identify alternative approaches.

Here we demonstrate that ligand-induced ERBB4 homodimers do not cause IL3-indpendent proliferation of 32D cells. In contrast, ligand-induced heterodimers of ERBB4 and ERBB2 do enable IL3 independence (**Table 11**). Therefore, we could generate a population of 32D cells that express *ERBB2* along with the entire set of 40 *ERBB4* mutant alleles found in *BRAF* WT melanomas. We could select for elevated signaling by ERBB4-ERBB2 heterodimers by propagating the bulk culture in the absence of IL3. We could identify the *ERBB4* mutant alleles that drive IL3 independence by deep sequencing the *ERBB4* cDNAs generated from the RNA of the bulk culture of IL3-independent 32D/ERBB2+ERBB4 cells. The relative frequency of each *ERBB4* mutant allele arising from this deep sequencing should correspond to the signaling activity of those *ERBB4* mutant alleles. In follow up experiments, we can also use a similar model system to determine whether specific *ERBB4* gain-of-function mutants cause increased ligand-dependent signaling, increased ligand-independent signaling, or both. Finally, should we demonstrate that *EGFR* is a driver of *BRAF* WT melanoma cell lines, we can use analogous approaches to explore the genetic interaction of *EGFR* with the 40 *ERBB4* mutant alleles found in *BRAF* WT melanomas.

### D. Our in silico analyses suggest strategies for treating ERBB4-dependent, BRAF WT melanomas

Here we demonstrate that, in the TCGA-SKCM *BRAF* WT melanoma dataset, *ERBB4* mutants are correlated with nonsynonymous mutations in a *RAS* gene or *NF1*. This surprising observation suggests that ERBB4 does not stimulate RAS signaling in *BRAF* WT melanomas. Instead, it appears that ERBB4 signaling cooperates with RAS signaling to drive *BRAF* WT melanomas.

We have previously demonstrated that the PI3K pathway is required for ERBB4-EGFR heterodimers to stimulate IL3 independent proliferation of BaF3 cells [35]. Likewise, a gain-of-function *ERBB4* mutant stimulates AKT phosphorylation in a human melanoma cell line [30].

Here we demonstrate that, in the TCGA-SKCM BRAF WT melanoma dataset, *ERBB4* mutations are somewhat inversely correlated with events predicted to cause ERBB4-independent activation of the PI3K pathway (mutation of *PIK3CA*, increased *PIK3CA* transcription, mutation of *PTEN*, or decrease in *PTEN* transcription). Thus, we have postulated that, in *BRAF* WT melanomas, *ERBB4* gain-of-function mutants stimulate PI3K signaling, which cooperates with elevated RAS signaling to drive the proliferation of these tumor cells.

Hence, we postulate that a suitable treatment for ERBB4-dependent, *BRAF* WT tumors would consist of a combination of a RAS pathway inhibitor and a PI3K pathway inhibitor. We believe that the most suitable RAS pathway inhibitor would be a MEK inhibitor (trametinib, binimetinib, selumetinib, or cobimetinib). The toxicity of the PI3K and mTOR inhibitors (alpelisib, sirolimus, everolimus, temsirolimus) may preclude the use of these drugs in this application. However, no specific ERBB4 inhibitors have been approved by the FDA. However, because ERBB4 mutants are likely to drive BRAF WT melanomas through signaling by ERBB4-EGFR or ERBB4-ERBB2 heterodimers, anti-EGFR drugs (gefitinib or cetuximab) or anti-ERBB2 drugs (lapatinib, trastuzumab, or pertuxumab) may be suitable. Future work is needed to evaluate these agents.

## Data Availability

All data produced in the present study are available upon reasonable request to the authors.

https://portal.gdc.cancer.gov/projects/TCGA-SKCM

